# An Economic Evaluation of a virtual Covid Ward in Leicester, Leicestershire, and Rutland

**DOI:** 10.1101/2022.06.27.22276736

**Authors:** Jim Swift, Noel O’Kelly, Chris Barker, Alex Woodward, Sudip Ghosh

## Abstract

**Objective:** The objective of this study was to demonstrate the impact of a virtual Covid—19 ward on NHS resource use.

**Methods:** Different methods were used to derive comparators for the observational data on acute length of stay versus the actual lengths of stay of 310 patients on acute wards and differences estimated. The resource use associated with delivering care in the virtual ward were collected on an ongoing basis.

**Results:** The virtual ward delivered estimated health care system savings of 1,103 bed days, £529,719 in net financial savings across two key groups of patients; those who had been on oxygen and required weaning off it while within the virtual ward and those not requiring oxygen therapy with less severe acute Covid disease. The costs of the intervention were 9.7% of the estimated gross savings and the mean net saving per patient was £1,709 in the base case without including the savings associated with a likely reduction in re-admissions. The 30-day re-admission rate was 2.9%, which was substantially beneath alternative comparative data. The mean cost of the intervention was £184.38 per patient.

**Conclusion:** The virtual ward delivered significant financial savings in both groups of patients, did so with a high degree of confidence, whilst doing so at a very low absolute and relative cost.

## Background

The Covid-19 pandemic placed substantial stress on the healthcare system in the UK and beyond diverting significant resources away from elective care and prioritising the acute care of unscheduled respiratory infections.

There are several indicators of the effects on the NHS in Leicester, Leicestershire and Rutland (LLR). At the end of March 2020, the University of Leicester Hospitals NHS Trust (UHL) was running at 53% of bed capacity (NHS England, Mar 2022). There was a need to increase space between beds to prevent infection and 19% of staff were absent because of illness or shielding. Acute hospitals are complex adaptive systems (NHS England, Mar 2022). It is to the credit of UHL managers and clinicians that 12 months later the hospital was running at 80% of its potential bed capacity and that further increased to 95% by the beginning of February 2022, (NHS England, Mar 2022).

The first main peak in infection was in April 2020 with health care systems seeking additional mechanically ventilated (MV) beds. In UHL, on 7^th^ April, 82% of occupied MV beds were occupied with patients suffering from Covid-19 respiratory syndromes (NHS England, Mar 2022). In January 2021, during the second wave of Covid-19 acute respiratory infections 87% of the UK’s critical care beds were occupied (NHS England, Mar 2022).

The NHS in LLR predicted a Covid-19 winter peak in 2020 and responded with several measures including the introduction of a virtual ward that commenced enrolling patients in November 2020 at the onset of the second wave of the Covid-19 pandemic in the UK. The two peaks of Covid-19 respiratory infections can be seen in figure 1 and there was very little difference between LLR and the rest of England in the scale of the impact of infections on hospital discharges. The primary objective of the development of the LLR Covid-19 virtual ward was the maintenance of acute capacity and the early safe discharge of care from the acute sector into an intermediate care setting supported with digital technology and The Leicester Partnership Trust (LPT) specialist respiratory team for people with Covid-19 respiratory infections. The standard operating procedure for the virtual ward is displayed on-line (SOP - LPT & UHL, Oct 2020).

**Figure 1.**
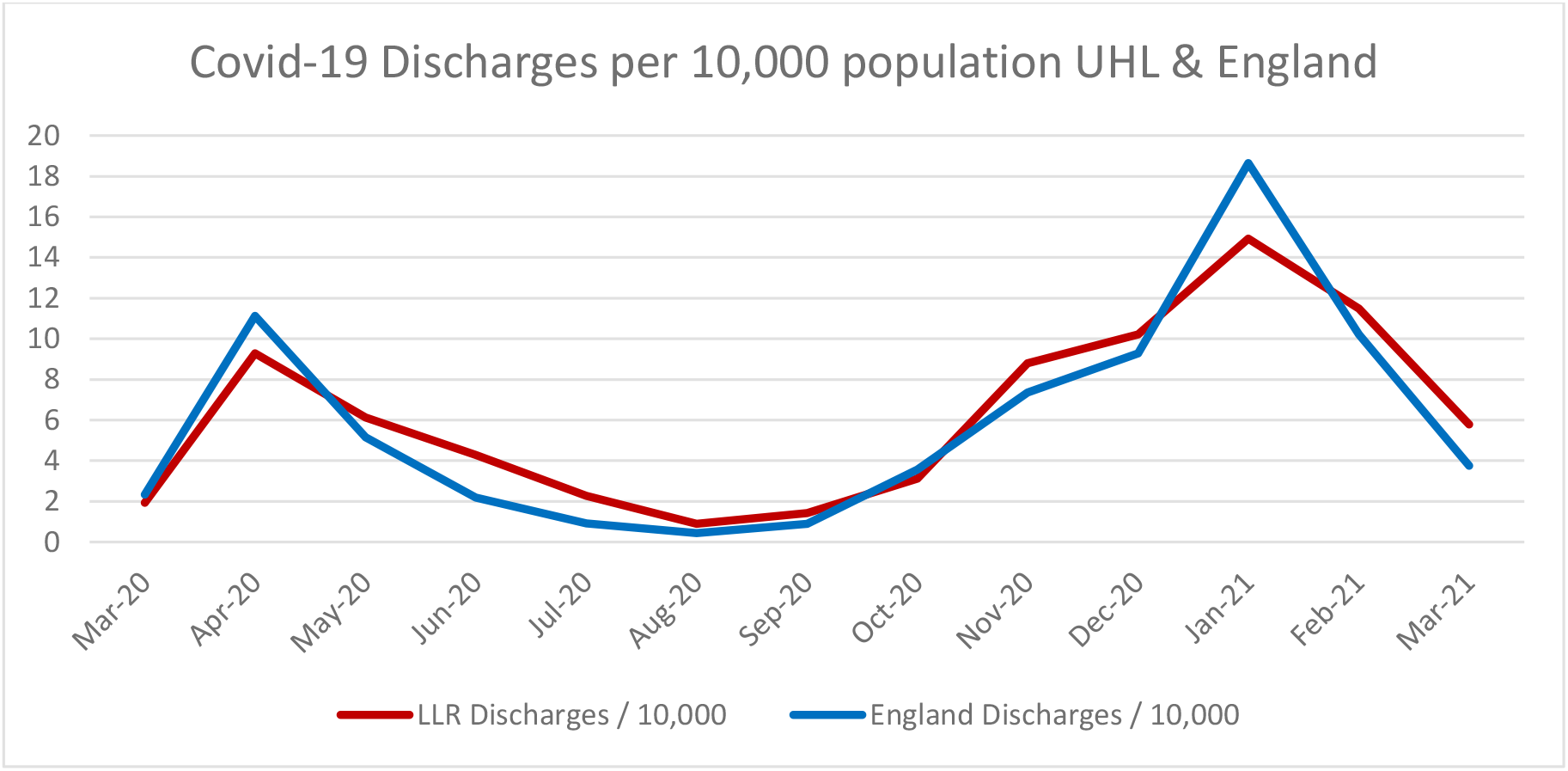
Covid-19 Discharges UHL & England per 10,000 population

## Methods

### Objective

The objective of this study was to demonstrate the impact of a virtual Covid ward on NHS resource use in secondary and intermediate care.

### Participants

310 patients discharged from UHL who had been admitted with Covid respiratory disease and were discharged into the virtual ward to either aid with oxygen weaning in their own home or discharged early to recover more fully at home and free up beds. The mean age of patients was 51.8 (median 56.0 years). 2.9% of patients were aged 80 years or older. The percentage of patients who were female was 40.6%. No ethnicity, co-morbidity or socio-economic status information was collected.

### Resource use and perspective

The data this study was based upon were observational and all patients discharged into the virtual ward had confirmed Covid-19 related respiratory disease. This is an economic analysis of a service evaluation. The CHEERS 2022 guidelines for economic evaluations were followed (Husereau D, 2022).

The perspective was that of the UK NHS. This was chosen as the perspective because any savings would be in the acute sector and costs in the intermediate care sector. It was not considered that

The comparator datasets were sourced from two NHS datasets, (NHS Digital, Aug 2021) and (NHS England, Mar 2022).

The resource use only considered acute hospital length of stay and the costs of running the virtual ward.

This analysis did not consider health effects beyond the resource implications associated with 30-day re-admissions. It was not considered that the virtual ward would deliver any change in patients’ long run health related quality of life or health outcomes.

The intervention was observed for 13 months, but no discounting of costs was performed because it barely extended beyond 12 months. The costs of bed days saved was sourced from UHL in November 2020 at the commencement of the intervention and all resource use were reported as 2020/2021 costs.

The analysis considered only the costs of the intervention and its’ comparator, remaining in an acute ward, and is a cost minimisation analysis. The outcome is reported in 2020 / 2021 pounds Sterling.

Acute resource use; lengths of stay (LOS), the cost of a day in a respiratory ward and re-admission within 30 days were sourced directly from UHL and the duration of LOS for patients discharged into the virtual ward were acquired from UHL [both correspondence Professor Ghosh S]. Staff unit costs were sourced from the PSSRU dataset for 20-21 (Jones K & Burns A, Dec 2021). The duration of the LOS for patients on oxygen not discharged into the virtual ward were acquired from UHL by LPT & NHS X, now part of the NHS Transformation Directorate and consultation durations and staff seniority were sourced from LPT [both correspondence Swift J]

### Comparison of acute ward stay versus imputed comparators

There were two distinctly different populations discharged into the virtual ward; those who had been on oxygen during their acute stay (n=31) awere subjected to an analysisnd discharged to be oxygen weaned at home in a virtual ward and those who were not on oxygen during their acute stay (n=279).

Those on O_2_ weaning were subjected to an analysis conducted by NHS X and LPT. NHS X personnel found that patients who accessed the virtual ward left hospital an average of 9.9 days before similar patients who had not accessed the virtual ward. The authors have accepted this external adjudication of resource use accurately reflected the savings made in bed days. The other group of patients were those who did not require oxygen during their stay in hospital.

In the analysis of the first 65 patients not on O_2_ (Swift J et al, 2021) who were discharged into the accelerated discharge programme were found to have left the virtual ward in 3.3 days, 2.2 days (40% relative reduction in LOS) earlier than controls who did not have the potential to access the virtual ward. The information on controls’ LOS was sourced immediately prior to the virtual ward’s introduction in November 2020. None of these patients had been in a critical care setting or had required oxygen. These patients were included in this analysis, as it is an analysis of all patients admitted into and discharged from the Covid virtual ward prior to end November 2021 and prior to the availability of newer medicines that reduced Covid illness severity.

### Data used to forecast LOS, patients not on O_2_ - Comparators

The length of stay in UHL was reported alongside median and mean length of stay against discharges between March 20 to March 21 with suspected and confirmed Covid-19 discharges (NHS Digital, Aug 2021). When the model moved the median LOS one month back in time the r^2^ improved from 0.48 to 0.94, see figure 2 for the comparison and appendix 1 for scatterplot. The relationship was linear.

**Figure 2.**
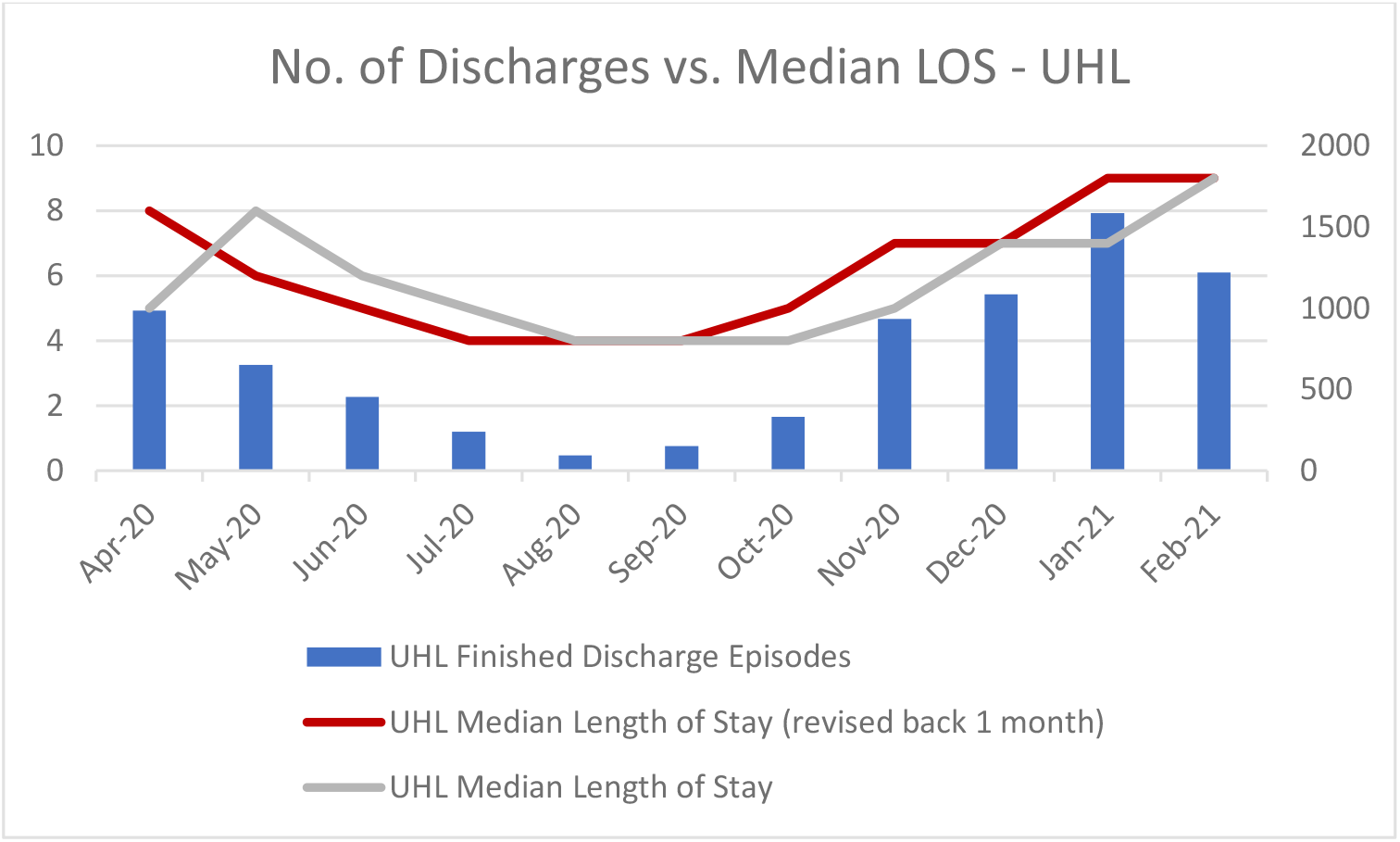
relationship Median LOS & No. of discharges

The original dataset (NHS Digital, Aug 2021) only ran to March 2021 and a different dataset with confirmed Covid-19 discharges was used but had no mean or median LOS data (NHS England, Mar 2022). This dataset was used as the basis for forecasting expected median LOS. The quality of the relationship (r^2^) in the adjusted dataset between discharge levels and median LOS deteriorated from 0.94 to 0.82 in the new dataset. However, the two datasets were highly correlated (r=0.98) and the relationship between the predictor (discharges) and predicted (median LOS) remained linear.

The mean LOS was not considered as a viable comparator because it substantially increased beyond the median because of the intensive resource use of patients who had extended stays to recover from critical illness and were further maintained on acute wards for recovery and to conduct oxygen weaning, see figure 3 for the difference between mean and median LOS. As this was a population that had not required oxygen, it was considered that the median LOS better reflected their estimated resource use.

**Figure 3.**
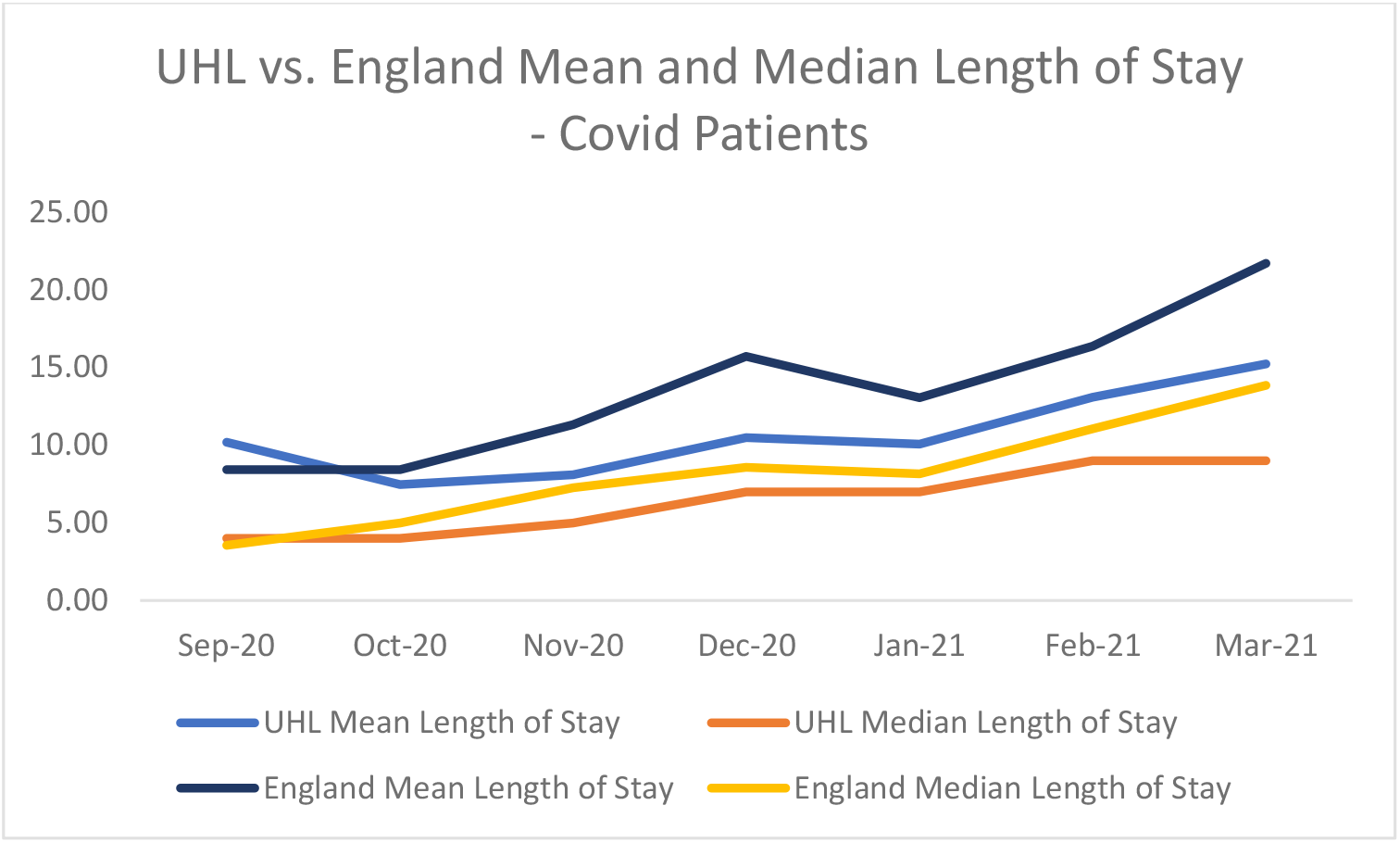
UHL Mean and median LOS: Patients with Covid-19

### Potential comparators to acute length of stay in non-O_2_ patients

Four additional comparators were developed to be used in a sensitivity analysis. Model 2 is the base case. Models 1-4 incorporated the impact that discharge levels had on the median length of stay. 196 patients who accessed the virtual ward (62.3% of all patients) accessed the virtual ward in the five months included in the original NHS Digital dataset; see figure 4. 62.3% of patients reported upon in the unadjusted ordinary least squares (OLS) model had an actual median LOS comparator not a modelled one. To create the better fit by pulling the median LOS data back one month meant that only 143 patients (46.1%) in the modified OLS and buckets datasets had median comparators not modelled ones. These were the best fitted datasets; median LOS vs. discharges (r^2^=0.85, 0.84, respectively). The fourth comparator was based on the adjusted median LOS but took account of the 10% longer LOS in the November patients than the median LOS. Model 5 was that used in the first analysis and was the length of stay of patients discharged prior to the inception of the virtual ward in November 2021. OLS modelling was considered most appropriate, as the data appeared to have a linear relationship, see appendix 1 for the median LOS and discharges scatterplots.

**Figure 4.**
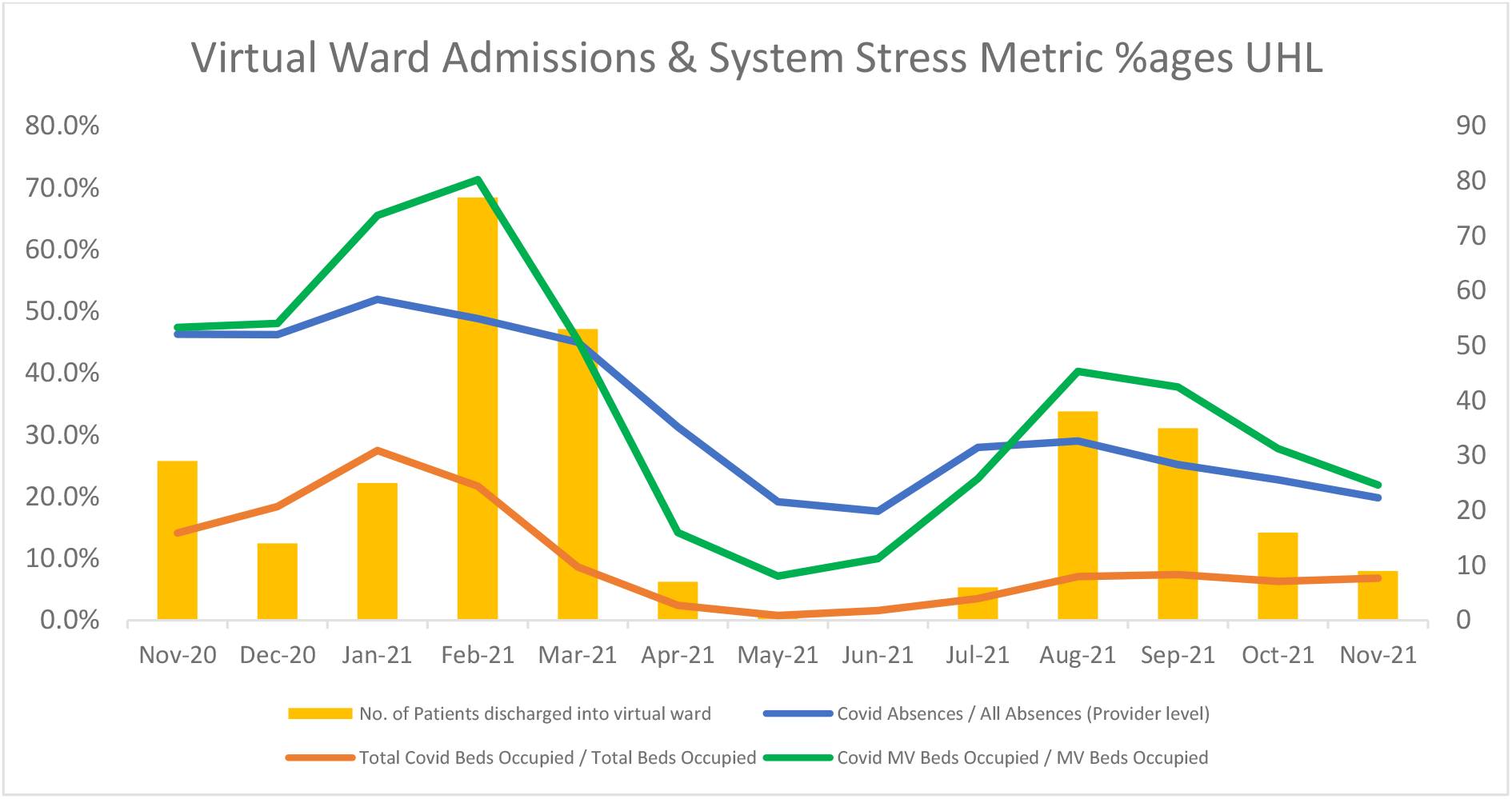
System demand and supply stressors: Beds, mechanically ventilated beds and absences

The five principal alternative methods used to determine comparators are described beneath:-

### Potential comparators

1. The ordinary least squares method was used to project the median length of stay forward using the number of monthly discharges as the independent variable (r^2^ =0.45, p<0.001). This did not take account of the better fit of moving the median data back one month.
2. The same as method 1 but took account of the better fit of moving the median data one month in arrears. This method provided the base case. (r^2^ =0.85, p<0.001).
3. Selecting buckets, where the median LOS were fitted to the boundaries of matching historic discharge levels and projected forward. Where boundaries overlapped the lower median LOS was allocated. The buckets projections were based upon the better fitting median LOS having been one month in arrears to retain the better fit, (r^2^ =0.84, p<0.001).
4. The original LOS data was 10% greater than the median LOS for November 2020. It was assumed that would persist in the modified OLS comparator.
5. The length of stay in the comparator group (no prior O_2_) in November 2020 was 5.5 days. It was assumed the mean LOS in November 2020 remained constant for comparable patients that did not access the virtual ward.

Three further methodologies were used to explore the above datasets.

6. A random number generator was used to generate numbers between the lowest and highest of the parameters (models 1-5) for each of the 279 patients and the data were compared to the acute LOS of non-O_2_ patients who accessed the Covid virtual ward. The random number generation was replicated 1,000 times and the mean taken for each patient.
7. The minimum value in the range for each patient was also explored.
8. The data were further explored by taking the better fitted modified OLS (base case) and buckets data and using it to add back the additional month of predictor data and reduce the predicted data by one month.

The one parameter that influenced the potential savings more than any other was the potential length of stay of any comparator to the acute length of stay prior to discharge into the virtual ward.

### Oxygen weaning patients - comparator

For the patients who had been on oxygen accessing the virtual ward an exercise was conducted by NHS X that demonstrated a reduced length of stay in the acute wards to have been 9.9 days. This was used as the comparator for all oxygen treated patients discharged into the virtual ward.

### Data Distribution

The data were not normally distributed in the datasets for either those who did not access oxygen and the oxygen weaned and the Wilcoxon Signed-Ranks Test for two paired samples was used to evaluate the data. The number of patients, (n=279 and n=331) were large enough to use normal approximation and a z-statistic was used.

Clinical specialist call duration and the costs of the digital technology as the two key input parameters related to costs of the intervention were varied by 50% for the sensitivity analysis.

### Analysis of the Length of Stay in the Virtual Ward

Patients in the virtual ward were monitored against a range of clinical criteria and a range of questions that related to their symptoms and self-assessed well-being. Patients were then classified by the algorithm into red, amber, and green (RAG ratings), which were highlighted on the LPT specialist respiratory clinicians’ desktop computers. Red alerts drove a clinical contact within 24 hours throughout the time patients were in the virtual ward. Amber in the first week meant a clinical contact, amber in the second week and thereafter did not require a contact. Green rated patients who had not been previously contacted were contacted during the second week.

RAG ratings were compared between the first and last days and the first 3 days and last 3 days. The distribution of the LOS and RAG ratings in the O_2_ patients were not normal and the Wilcoxon Signed-Ranks Test for two paired samples was used to evaluate the data. The LOS data was not normal for the acute and virtual wards, but the RAG ratings were normally distributed in the non-O_2_ patients, and a two tailed students t-test was used to compare the matched pairs.

All analyses were conducted in Microsoft Office Excel Analysis Toolpak.

## Results

The two different patient populations; oxygen weaning and non-O_2_ weaning had very different lengths of stay acutely and in the virtual ward reflecting the severity of their Covid illness and the time taken to reduce their oxygen dose. For patients not on oxygen, the mean acute LOS was 4.2 days (median 4.1, standard deviation σ 2.1). For patients on oxygen weaning in the virtual ward, the mean acute LOS was 13.7 days (median 12.4, σ 4.5).

In the 310 patients the mean acute LOS was 5.1 days, (median 4.6, σ 3.7). 90% of patients had not required oxygen prior to discharge into the virtual ward.

The number of patients discharged into the virtual ward tended to reflect the overall demand in the acute system and capacity issues with perhaps an exception at the introduction of the intervention. This may have reflected a degree of clinical conservatism with an unknown entity, see figure 4. The number of discharges into the Covid ward (yellow bars in figure 4) corresponds with key system stressors rising and falling; Covid absences over all absences UHL, Covid-19 beds occupied over all beds and Covid-19 occupied mechanically ventilated (MV) beds over all MV beds.

### Non-O2 Patients

#### Acute LOS

The mean acute ward LOS of the 279 patients accessing the virtual ward who had not been on oxygen was 4.2 days. Table one beneath demonstrates the differences between effect sizes of the different potential comparators described in the methods section with the actual mean lengths of stay in the acute wards prior to discharge into the virtual ward. All were applied as comparators to the lengths of stay in UHL with the second option being the base case. These data were then used to develop a further comparator using random numbers between the lowest and highest median LOS for each patient.

The actual mean LOS in acute wards in UHL for those who accessed the virtual ward is compared to the UHL median LOS and discharge numbers in Figure 4. Figure 4 shows the gulf between the median length of stay and actual LOS in patients discharged into the Covid-19 virtual ward as being much larger than the forecasted gulf. The mean LOS in those discharged into the virtual ward tracks well beneath the median acute LOS. The median LOS data presented in figure 4 stopped at March 2021. The data were unavailable for the mean and median LOS thereafter and were estimated. The methods for estimation were described in the methods section and the table of results are displayed beneath in Table one.

The data presented in table 1 are graphically represented beneath in figure 4. The left hand Y axis is the median UHL LOS and the right hand Y axis is the number of UHL Covid-19 discharges. All five of the potential comparators were plotted against the actual LOS pre-discharge into the virtual ward shown in a red hashed line. There were no discharges into the virtual ward in June 2021, hence the gap in the actual LOS pre-discharge into the virtual ward shown in Figure 5.

**Table 1.** Comparison with Acute Length of Stay Prior to Discharge into Virtual Ward (Non-O_2_ Weaning)

| Comparator Table | Ordinary Least Squares 1 | Modified Ordinary Least Squares 2 | Buckets 3 | Modified Ordinary Least Squares +10% 4 | Constant 5.5 days 5 |
| --- | --- | --- | --- | --- | --- |
| W min | T=2915.5 | T=2463 | T=3030.5 | T=1401.5 | T=6552.5 |
| z-statistic | z=12.317 | z=12.652 | z=12.232 | z=-13.439 | z=9.621 |
| $\alpha=0.01$ (zc) | zc=2.58 | zc=2.58 | zc=2.58 | zc=2.58 | zc=2.58 |
| Test p value | p<0.01 | p<0.01 | p<0.01 | p<0.01 | p<0.01 |
| Mean acute LOS days | 6.99 | 7.05 | 6.91 | 7.76 | 5.50 |
| Mean Absolute $\Delta$ | 2.80 | 2.86 | 2.71 | 3.56 | 1.30 |
| Mean Relative $\Delta$ | 40.0% | 40.5% | 39.2% | 45.9% | 23.7% |
| Data fit (r<sup>2</sup>) | 0.45 | 0.85 | 0.84 | 0.85 | N/A |
| Linear Equation | $y = 0.0073x + 3.7183$ | $y = 0.0083x + 3.8526$ | $y = 0.0086x + 3.6234$ | $y = 0.0091x + 4.2378$ | $y = 5.5$ |

**Figure 5.**
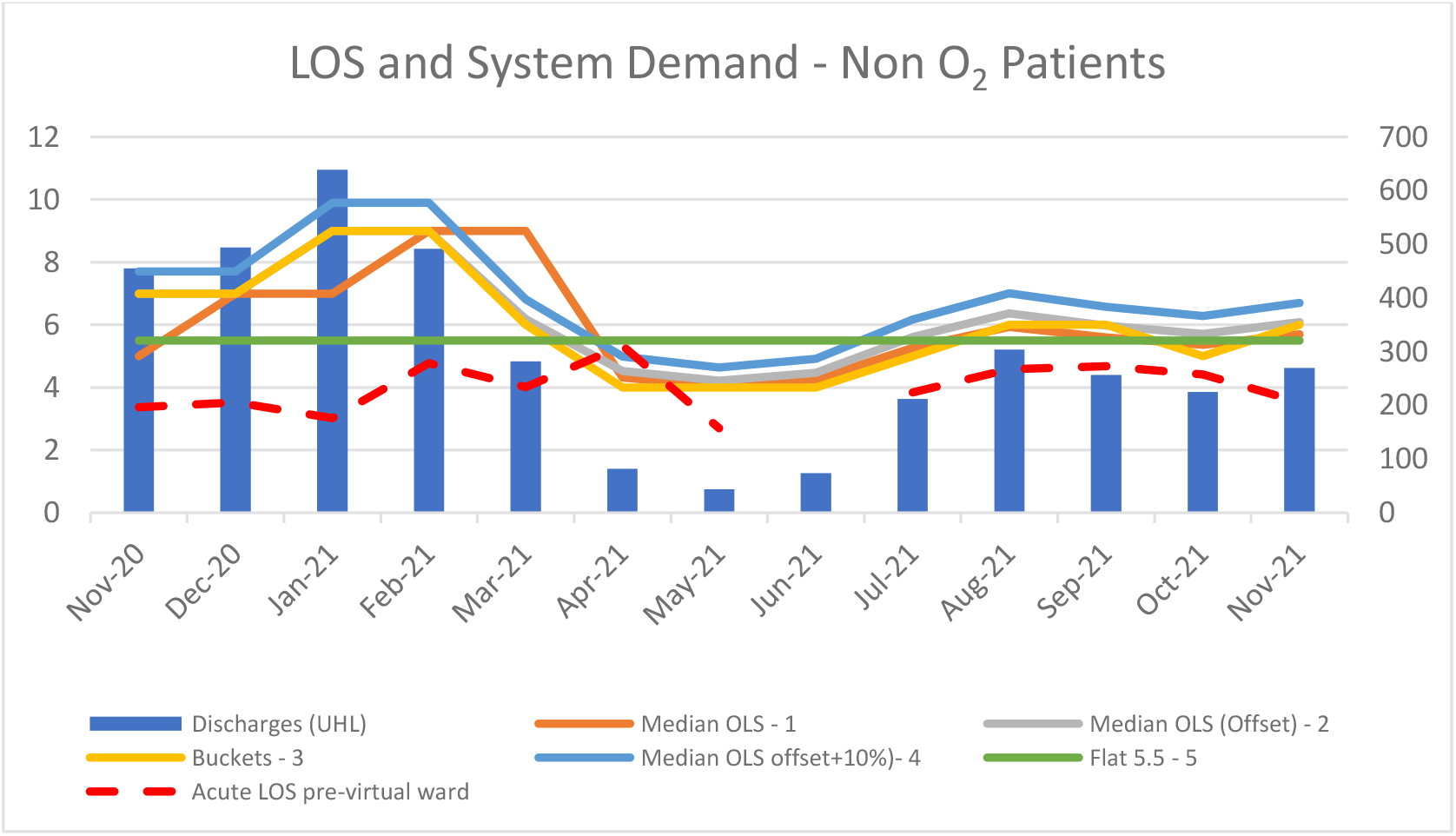
Median acute LOS comparators versus discharges by month

**Table 2.** Comparison with Acute Length of Stay Prior to Discharge into Virtual Ward (O_2_ Weaning)

| Comparator Table | Patient days on acute ward pre discharge to virtual ward | Comparator |
| --- | --- | --- |
| W min |  | T=0 |
| z-statistic |  | z=4.86 |
| $\alpha=0.01$ (zc) | | zc=2.58 |
| Test p value |  | p<0.01 |
| Mean acute LOS | 13.27 | 23.17 |
| Mean Absolute $\Delta$ | | 9.90 |
| Mean Relative $\Delta$ | | 42.7% |

**Table 3a.** Staff unit costs

| Staff costs | Unit Cost £ |
| --- | --- |
| Band 6 nurse / physio per hour | £54 |
| Band 7 nurse / physio per hour | £65 |

**Table 3b.** Staff time per consultation

| Consultation duration | Time in minutes |
| --- | --- |
| Time for consultation 1 | 20 |
| Time for consultation 2 | 7.5 |
| Total time | 27.5 |

**Table 3c.** Staff costs per consultation

| Nurse / Physio call costs – RAG | Costs per virtual consultation |
| --- | --- |
| Band 6 | £24.75 |
| Band 7 | £29.79 |

**Table 3d.** Costs of digital technology per day

| Costs of CliniTouch Vie | CliniTouch Vie per diem |
| --- | --- |
| Rate | £3 |

**Table 4a.** Total & mean costs of specialist respiratory nurse / physio consultations

| Specialist nurse / physio calls | No. | Cost |
| --- | --- | --- |
| Reds (total population 310) | 909 | £27,081 |
| Ambers (week 1 only) | 438 | £13,049 |
| Greens not contacted in week 1 (week 2 only) | 61 | £1,817 |
| Total calls | 1,408 | £41,947 |
| Per patient |  | £135.31 |

To aid with model conservatism, all contacts were assumed to have been made by band 7 specialists.

**Table 4b.**
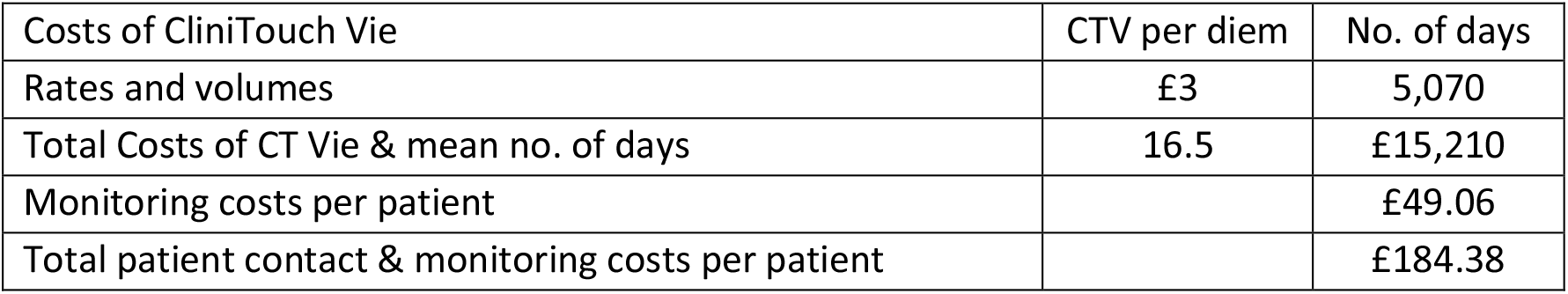
Costs of digital technology use in the virtual ward & mean virtual ward costs

**Table 5a.** Breakdown in Resource Use in the virtual ward in units

| Resource Use | Number of Patients | Reduction in LOS (Days) | No. of days in VW | Red Alerts | Amber Alerts Week1 | Green Alerts Week2 |
| --- | --- | --- | --- | --- | --- | --- |
| O2 | 31 | 306.9 | 1,344 | 313 | 44 | 5 |
| Non-O2* | 279 | 796.3 | 3,726 | 596 | 394 | 56 |
| Combined | 310 | 1103.2 | 5,070 | 909 | 438 | 61 |

\*Using the OLS adjusted model as the comparator in the base case for resource use in the non-O<sub>2</sub> population

**Table 5b.** Breakdown in Resource Use in The Virtual Ward in £s

| Resource Use | Reduction in LOS (Days) | Costs of Digital | Costs of Virtual Consults | Total Costs | Net Savings |
| --- | --- | --- | --- | --- | --- |
| O2 | £163,271 | £4,032 | £10,785 | £14,817 | £148,454 |
| *Non-O2 | £423,605 | £11,178 | £31,162 | £42,340 | £381,265 |
| Combined | £586,876 | £15,210 | £41,947 | £57,157 | £529,719 |

\*Using the OLS adjusted model as the comparator in the base case for resource use in the non-O<sub>2</sub> population

**Table 5c.** Breakdown in Resource Use in The Virtual Ward in £s per patient

| Mean Resource Use per Patient | Reduction in LOS (Days) | Costs of Digital | Costs of Virtual Consults | Total Costs | Net Savings |
| --- | --- | --- | --- | --- | --- |
| O2 | £5,267 | £130 | £348 | £478 | £4,789 |
| *Non-O2 | £1,518 | £40 | £112 | £152 | £1,367 |
| Combined | £1,893 | £49 | £135 | £184 | £1,709 |

\*Using the OLS adjusted model as the comparator as the base case for resource use in the non-O<sub>2</sub> population

**Table 6.** 30-day readmissions costs

| Re-admissions Quantity & Costs | No. | Cost |
| --- | --- | --- |
| Re-admissions within 30 days ** | 9 | £3,676 |
| Total re-admissions rate and costs | 2.9% | £33,085 |
| Expected re-admissions rate and costs | 7.1% | £99,951 |
| Per patient |  | -£190.58 |

\*\* it was assumed that the length of stay was 7.05 days, as per the adjusted OLS model.

The data showed a substantially better fit by moving the median length of stay back one month in time. When the modified values were inserted back into the original time series giving an additional predictor month in March 2021 for the buckets and modified OLS data (the base case), the mean change in LOS predicted increased by 0.03 and 0.06 days respectively. Whilst it made more intuitive sense to replace values in a time series where a value was skewed one month into the future, in practical terms, it made no real difference to the findings. The original values were retained as they were lower, retaining the conservatism of the findings.

An analysis of length of stay of patients admitted to hospital using three different methods to establish a projected overall length of stay suggested it would range from 8.0 to 9.1 days for patients not admitted to an ICU (Vekaria B, 2021). All estimates used as comparators in the non-O_2_ weaning group fell below this range. The highest single mean value was 7.76 in the potential comparators used and the base case LOS used in the resource use calculations was 7.05 days. Further, the mean reduction in LOS between November and March was 3.7 days in actual median data versus actual LOS data whereas the fall in the difference in predicted median LOS versus actual LOS fell to 1.1.

When the lowest potential comparator for each of the first three methods, which excluded the highest LOS value was compared against the actual length of stay for each patient, the mean LOS was reduced to 6.5 days and the gain was reduced to 2.3 days. When all five potential methods were considered and the minimum value only was taken for the comparator LOS, the LOS dropped to 5.4 days, which meant the potential difference was reduced to 1.2 days. The mean of all five methodologies with the appropriate upper and lower limits with 1,000 random simulations was 6.5, which was a reduction of 2.3 days.

The difference between the actual acute median LOS data (mean 8.0 days) and the median LOS projected data (adjusted OLS = 5.3 days) was 2.7 days per stay. Whilst this may look like a mis-specification. The projected median LOS data reflected the reduced discharges that followed the second peak in the Covid-19 pandemic. In February and March 2021 some 7.1% of all acute discharges in UHL were admitted to the virtual ward. The mean and median LOS in England increased as discharges also increased in a similar pattern to that of UHL.

The mean virtual ward LOS was 13.4 days for patients who had not been on oxygen, (median 15, σ 4.9).

### Patients on Oxygen

The mean virtual ward LOS for the 31 patients who had been on oxygen was 46.3 days, (median 37, σ 32.4).

### Resource Use in The Virtual Ward

There were 310 patients discharged into the virtual ward.

The bed days estimated to be saved were 306.9 from the 31 patients who were oxygen weaned in the virtual ward.

The range of bed days estimated to be saved from the 279 patients who had not been on oxygen varied from the lowest estimate of 363.8 (vs. method 5 - constant 5.5 days comparator), to the highest which was 994.3 (vs. method 4 - modified OLS+10%). The intervention reduced bed days by 779.9 and 756.3 days versus methods 1 (OLS) and 3 (buckets) respectively. Method 2 was the base case.

The costs of running the virtual ward were clear and reflected the resource use associated with patient duration in the virtual ward and the demand on clinician time directly related to the indicator of patient symptoms and wellbeing, the RAG rating.

### Resource Use Unit Costs

The 30-day re-admission rate was lower than that expected looking at the literature on 30-day re-admissions. A systematic review that regarded the question (Ramzi ZS et al., 2022) found that the mean rate of 30-day readmissions was 10.34%. Of the studies conducted in the UK, three reported on 30-day readmission rates, (Chaudhry, 2021) found 10.2% readmissions, however, the study included seriously ill patients whilst only 10% of the patients included in this study were seriously ill requiring critical care and or oxygen. (Mooney, 2021) and (Spence, 2021) reported 11.3% and 17.1% readmissions respectively, but their findings were in elderly patients. The patient population in this analysis had a mean age of 54.6 years and only 2.9% of patients were aged 80 years or older. These studies were not considered representative. A study conducted in a Turkish tertiary centre (UyaroĞlu et al, 2021) found that 7.1% of patients with mild or moderate Covid disease, i.e. in a broadly similar population were readmitted within 30 days. If this study was considered representative, there would have been a 59.2% reduction in 30-day re-admissions in a naïve comparison with the most appropriate and conservative evidence-based comparator.

None of the 31 patients discharged who had been on oxygen were re-admitted within 30 days. All nine of the re-admissions were in those who had not accessed oxygen during their acute stay. 90% of patients discharged from UHL into the virtual ward had not been on oxygen.

The mean unit costs per patient stay in the virtual ward varied depending upon how the potential gain in 30-day re-admissions was treated. If the saving was ignored the mean unit cost was £184.38. If it was allocated as a saving the mean unit cost per patient in the virtual ward was -£9.02 in the base case. The cost of a bed day in a respiratory ward in University Hospitals Leicester in November 2020 was £532. This cost was applied to the number of bed days released in the virtual ward between November 2020 and November 2021 (inclusive). No adjustment was made for inflation.

The cost of an average stay in the virtual ward (£184.38) was 34.7% of the cost of a bed day (£532).

### Virtual Ward Activity

310 patients accessed the virtual ward. It would be expected that after an acute infection that people would generally recover. The resource use implications of virtual ward activity have been explored as have re-admissions. The trends towards recovery can be seen in the RAG ratings over time.

For those on oxygen weaning the percentage of patients with a red day on their first day in the virtual ward was reduced from 45% to 14% on their last day. For those who had not been on oxygen the percentage was reduced from 21% to 13%. The percentage of patients who had a red RAG rating on any of the first or last 3 days reduced from 55% to 31% and 40% to 19% respectively. All results were highly statistically significant at p<0.01 except for the O2 weaning first three days versus last three days (p=0.024). The data for the O2 weaning was not normally distributed and a Wilcoxon Signed Rank Test was used. The data for the patients who had not been on oxygen was normally distributed and a students t test was used.

79% of virtual ward participants required a specialist health care professional contact in their first three days in the O2 weaning group having either a red or amber RAG rating. In the patients who had not been on oxygen this was still a relatively high 67%. Overall, 68% of participants required a specialist respiratory contact in their first three days in the virtual ward, which supported the contention that these patients had a reduced health status and were unfit to be discharged home without additional clinical support.

43 patients had a RAG rating of two reds and an amber or three reds in their first three days. If these patients had been kept in an acute ward for observation for these three days, they would have consumed more resources (£68,628) than the cost of the virtual ward for all 310 patients (£57,157).

### Sensitivity Analysis

The duration of consultations were varied by 50%, as was the cost of the digital technology, CliniTouch Vie (Spirit Health). These two elements were the key drivers of the costs of the virtual ward. When the impact of both was combined and the cost of the virtual ward was increased by 50%, the overall impact on costs when both were simultaneously applied was an increase from £184.38 to £276.56. However, as the costs of the virtual ward were only 9.7% of the gross savings, this had a relatively insignificant effect on the overall savings, which were reduced from £1,709 in the base case to £1,617.

The key parameter which influenced the outcome most was the acute length of stay. The modified OLS method provided the base case; four other plausible alternatives, as described in the methods section were considered and the results shown in tables 7a and 7b.

**Table 7a.** Scenario 1 where estimated readmission savings were unallocated

| Summary of v ward resource use | modified OLS | OLS | Buckets | Mod OLS +10% | Flat 5.5 days | All |
| --- | --- | --- | --- | --- | --- | --- |
| Costs excluding readmissions savings | -£57,157 | -£57,157 | -£57,157 | -£57,157 | -£57,157 | -£57,157 |
| Non O<sub>2</sub> patients | £423,605 | £414,699 | £402,825 | £528,956 | £193,542 | £336,032 |
| O<sub>2</sub> weaning patients | £163,271 | £163,271 | £163,271 | £163,271 | £163,271 | £163,271 |
| Total net savings | £529,719 | £520,813 | £508,939 | £635,070 | £299,656 | £442,147 |
| Per patient saving | £1,709 | £1,680 | £1,642 | £2,049 | £967 | £1,426 |

**Table 7b.**
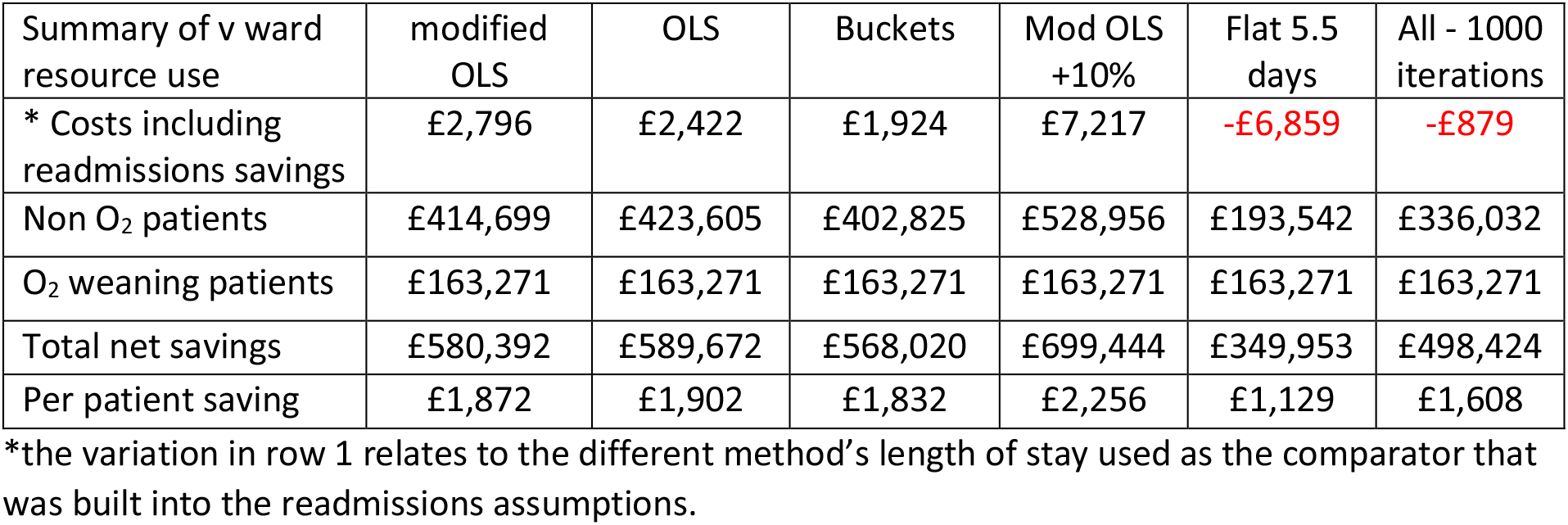
Scenario 1 where estimated readmission savings were allocated

It can be seen in table 7a that the costs of delivering the virtual ward were £57,157 and the savings for the patients who had been on oxygen were £163,271. The base case savings where no re-admissions savings were allocated, and net of costs were £529,719 or £1,709 per patient. The random model with 1,000 iterations between the lowest and highest value of all five methods delivered £442,147 in net savings or £1,426 per patient.

The gross saving per patient in the base case excluding any potential savings made in re-admissions was £1,893 with costs of £184.38 (9.7%) per patient.

The lowest daily length of stay across the variables used as parameters was also considered. In this scenario the reduction in length of stay was reduced to 1.2 days. Even at that level the gross savings would have been £337,660. When the costs of the virtual ward intervention were increased by 50% as per the sensitivity analysis, the savings whilst somewhat diminished would have been £251,925 or £812.66 per patient.

## Results summary

The greatest area of uncertainty was the saving in bed days in those who were not on oxygen prior to their discharge into the virtual ward.

Background evidence, as demonstrated in figure 3 of lower median LOS in UHL versus England and a flattening of the median UHL LOS curve, which corresponded with peak patient discharges into the virtual ward contrasted with the continued growth in median LOS in England. 106 patients (38%) of all non-O_2_ discharges into the virtual ward were in these two months. It is noteworthy that the mean curve did not track the flattening of the median one in UHL, which reflected the small absolute number of discharges of oxygen weaning patients into the virtual ward at that time. The change in the slope of the median LOS curve in figure 3 supports the face validity of the results that bed days were reduced after the introduction of the virtual ward.

That patients left hospital early is not really in any doubt, it is the extent to which they did that is. However, the proximity of the first three methods; OLS, modified OLS and buckets did seem to suggest there was a reduction of around 2.8 days (2.71-2.86) in patients who had not been supported on oxygen’s acute length of stay. The mean reductions in bed days ranged from 3.43 to 3.56, a 40.2% to 41.1% relative reduction in bed days in the same groups once the oxygen weaning patients were included. The reasons why the modelled comparators showed such a low difference between the median LOS and the pre-discharge into the virtual ward was related to the linear relationship between demand and median LOS. The buckets method is a linear one as it is also dependent upon historic discharge levels versus median stay and forecasts based on that; albeit it is more conservative as the lowest values were used when boundaries overlapped. The virtual ward was clearly cost saving for patients on oxygen and by a very substantial margin of 9.9 days per patient and a reduction of 42.7% in bed days.

The costs of the virtual ward versus the savings estimated in the cost of bed days (excluding any potential readmission savings) were 9.1% (O2 weaning) and 10.0% for those who hadn’t accessed oxygen in hospital. The absolute costs were £477.95 and £151.76 respectively. The higher virtual ward costs for those discharged for oxygen weaning reflected a higher mean contact rate with the specialist respiratory clinicians 10.1 versus 2.1 red days and longer stays in the virtual ward, 43.4 versus 13.4 days. The net savings were also substantially greater in the oxygen treated patients; £4,789 versus £1,367. The total costs as a percentage of gross estimated savings were 9.7%.

## Discussion

The absence of a definitive comparator renders a degree of uncertainty around the savings, which is why different potential comparators were explored so extensively in the sensitivity analysis. The virtual ward was a pragmatic response to a pandemic with a predicted surge in winter pressures forecast to be driven primarily by Covid-19 respiratory infections. However, the estimates associated with the comparators’ resource use always erred on the conservative. 7.1% of all UHL discharges in February / March 2021 were discharged early into the virtual ward thereby reducing the denominator. It is highly likely that the savings were underestimated.

The virtual ward intervention saw a substantial reduction in 30-day readmissions versus other observational data in a naive comparison, which is not proof of either an effect or safety but is a strong positive signal.

The UK has a crisis in demand for elective care driven primarily by the shortfall in supply during the two years of the pandemic in an ageing population.

The Covid-19 pandemic saw the use of remote and digital care rise as a response to the need to provide care remotely where possible, but in the Covid-19 virtual ward it was more a planned response to an anticipated surge in Covid-19 related demand and constrained supply. The UK NHS has banked on digital to prove a successful vehicle to reduce acute admissions and re-admissions by helping keep people well, keeping them out of hospital, supporting their discharge and freeing up much needed elective capacity to reduce the number of patients waiting and the duration of their waiting times.

The Covid-19 virtual ward intervention achieved its primary goal of increasing acute capacity and was used most when capacity was most pressured. It also reduced health care resource use.

There is an almost universal geographical availability of specialist community respiratory teams competent to manage people discharged from acute care early in the UK health care system. The almost infinite scalability of the technology means there is little technological barrier to wider use. 83% of people between 55 and 64 owned a smartphone in 2021, an increase of 13% from 2020 (O’Dea S, 2021). The mean age of patients accessing the virtual ward was 52. The possibility of new variants, the low financial and safety risk to health systems coupled with the scalability of this and other digital technologies, widespread societal digital literacy, and access to smartphones, except perhaps in the very oldest segments of the least affluent in society, many of whom will reside in care homes and carers could monitor their wellbeing has meant that there is an opportunity to be more prepared for future pandemics.

The combined efforts of everyone involved in the design and delivery of the virtual ward demonstrated the willingness of people to work across systems within the NHS; all three LLR CCGs; NHS Leicester City, NHS East Leicestershire and Rutland, NHS West Leicestershire, University Hospitals Leicester NHS Trust, The Leicestershire Partnership NHS Trust and with the private sector partner Spirit Health who delivered the digital technology to deliver a successful intervention that helped reduce the demand pressures in the acute system. It should not be underestimated the extent to which Covid-19 fostered a willingness to co-operate and how important that was.

## Conclusion

The resources used in the virtual ward were lower than the comparators and 1,064 - 1,103 bed days were estimated to have been saved. The financial savings were estimated to be between £508,939 and £529,719 with an average saving of between £1,642 and £1,709 per patient when methods 1-3 were used.

The costs of the virtual ward were relatively low compared to the savings. Overall, a cost to savings ratio of 9.7% when the modelling was conservative at every turn suggests the financial risk of this not being cost saving is very low to health care systems.

## Supporting information

Appendix 1 Scatterplots

CHEERS 22

## Data Availability

All data produced in the present study are available upon reasonable request to the authors

## Author Declarations

Ethical committee approval was not necessary as this was an economic analysis of a service evaluation.

## Thanks

The authors would like to thank Zoe Harris from the NHS Transformation Directorate who contributed significantly to the first paper and shared useful information for this paper and Irene Valero-Sanchez, respiratory consultant from University Hospitals Leicester NHS Trust for helping develop and steer the project.

