## Appendix 1 Scatterplots for "An Economic Evaluation of a virtual Covid Ward in Leicester, Leicestershire, and Rutland"

### *Ordinary Least Squares*

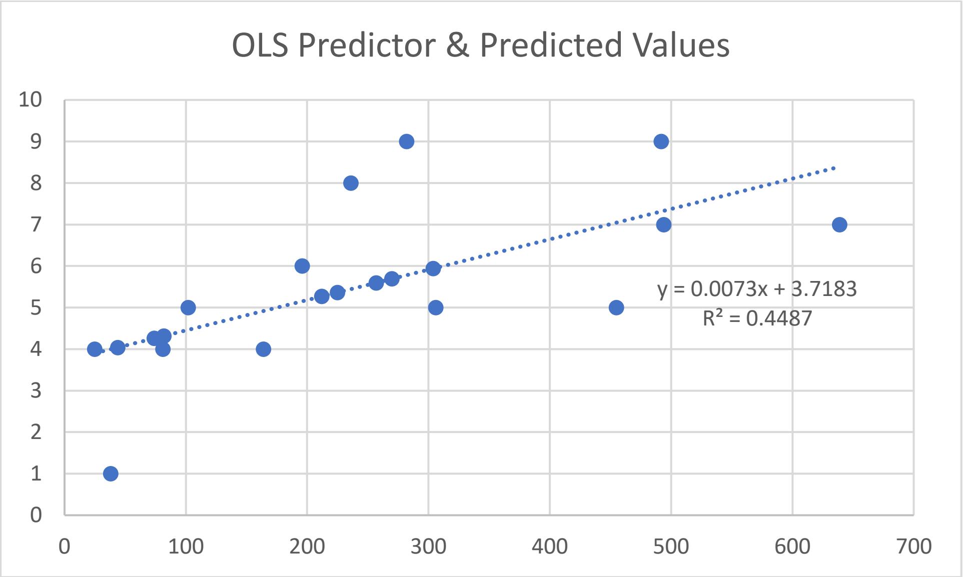


### *Ordinary Least Squares (modified by moving the median LOS one month back in time)*

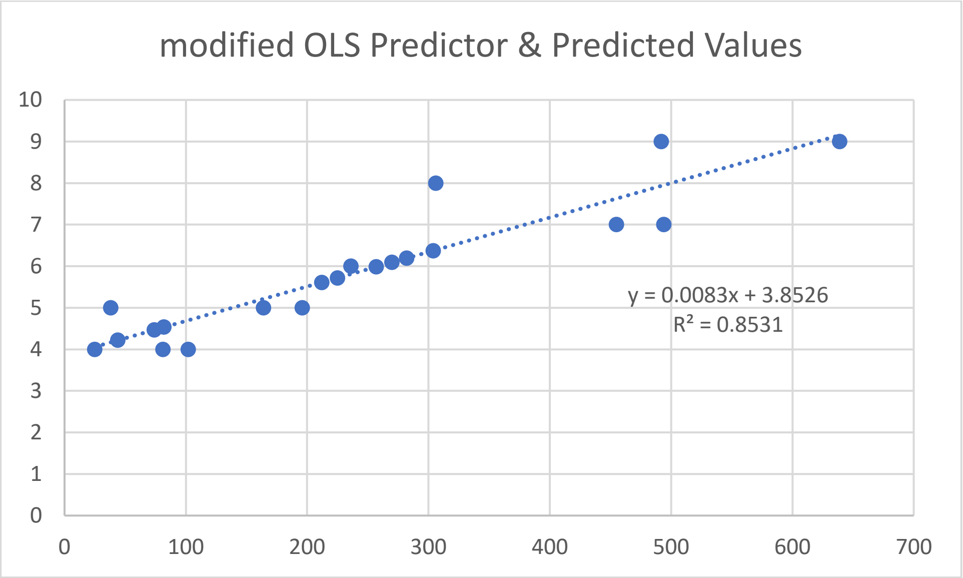

*Buckets*

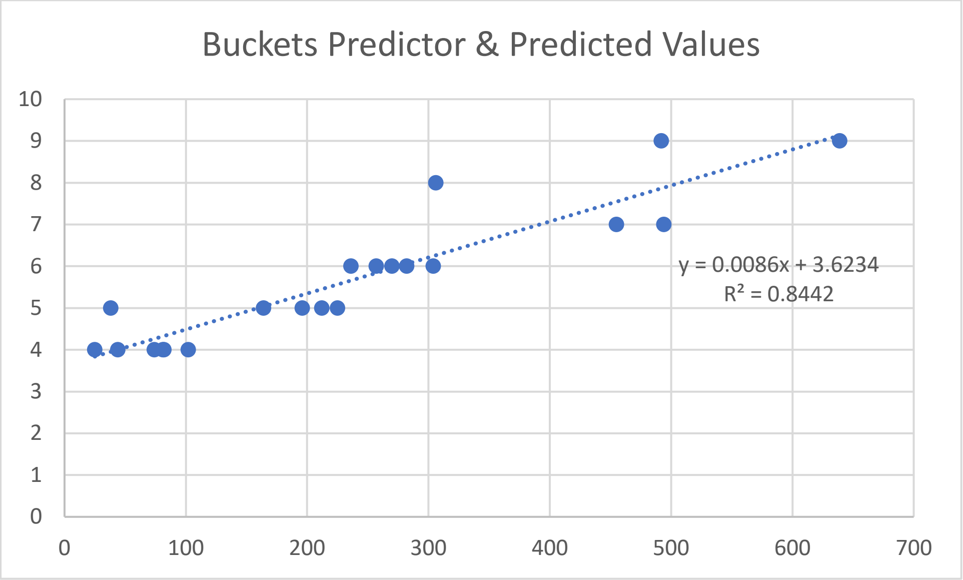


*Ordinary Least Squares (modified by moving the median LOS one month back in time) plus 10%*

This dataset reflects the 10% longer LOS in the population discharged prior to the introduction of the virtual ward over the median LOS in November 2020.

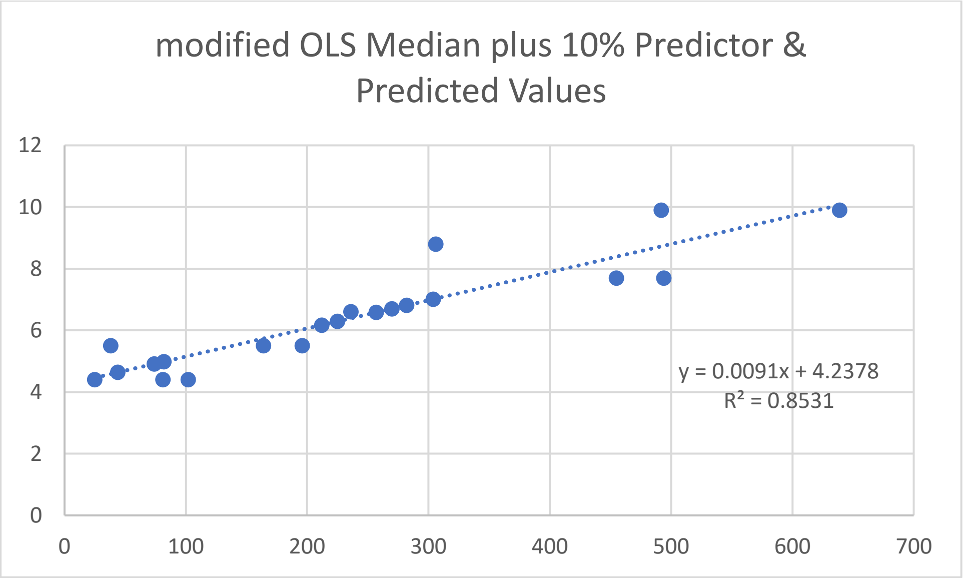
