## Supplementary material for "An Economic Evaluation of a virtual Covid Ward in Leicester, Leicestershire, and Rutland": CHEERS 22

| | | | |
|---|---|---|---|
| | CHEERS 22 | Title: An Economic Evaluation of a virtual Covid Ward in Leicester, Leicestershire, and Rutland | |
| Title | Title | Identify the study as an economic evaluation and specify the interventions being compared. | Pages: Title (1) and page 3 |
| Abstract | Abstract | Provide a structured summary that highlights context, key methods, results, and alternative analyses. | Pages: Abstract (1) |
| Introduction | Background and objectives | Give the context for the study, the study question, and its practical relevance for decision making in policy or practice. | Pages: 2, 15 |
| Methods | Health economic analysis plan | Indicate whether a health economic analysis plan was developed and where available. | N/A as simple CMA |
| Methods | Study population | Describe characteristics of the study population (such as age range, demographics, socioeconomic, or clinical characteristics). | Pages: 3, 12 |
| Methods | Setting and location | Provide relevant contextual information that may influence findings. | Pages: 2,3 |
| Methods | Comparators | Describe the interventions or strategies being compared and why chosen. | Pages: 3, 4, 5, 6 |
| Methods | Perspective | State the perspective(s) adopted by the study and why chosen. | Page: 3 |
| Methods | Time horizon | State the time horizon for the study and why appropriate. | Page: 3 |
| Methods | Discount rate | Report the discount rate(s) and reason chosen. | Page: 3 |
| Methods | Selection of outcomes | Describe what outcomes were used as the measure(s) of benefit(s) and harm(s). | Page: 3 |
| Methods | Measurement of outcomes | Describe how outcomes used to capture benefit(s) and harm(s) were measured. | Page: 3 |
| Methods | Valuation of outcomes | Describe the population and methods used to measure and value outcomes. | Pages: 3, 4 |

| | | | |
|---|---|---|---|
| Methods | Measurement and valuation of resources and costs | Describe how costs were valued. | Pages: 3, 9, 10, 11 |
| Methods | Currency, price date, and conversion | Report the dates of the estimated resource quantities and unit costs, plus the currency and year of conversion. | Page: 3 |
| Methods | Rationale and description of model | If modelling is used, describe in detail and why used. Report if the model is publicly available and where it can be accessed. | Pages: 3, 4, 5, 6 |
| Methods | Analytics and assumptions | Describe any methods for analysing or statistically transforming data, any extrapolation methods, and approaches for validating any model used. | Pages: 3, 4, 5, 6 |
| Methods | Characterising heterogeneity | Describe any methods used for estimating how the results of the study vary for subgroups. | Pages: 4, 5, 6 |
| Methods | Characterising distributional effects | Describe how impacts are distributed across different individuals or adjustments made to reflect priority populations. | N/A |
| Methods | Characterising uncertainty | Describe methods to characterise any sources of uncertainty in the analysis. | Pages: 4, 5, 6, 8, 9 |
| Methods | Approach to engagement with patients and others affected by the study | Describe any approaches to engage patients or service recipients, the general public, communities, or stakeholders (such as clinicians or payers) in the design of the study. | N/A |
| Results | Study parameters | Report all analytic inputs (such as values, ranges, references) including uncertainty or distributional assumptions. | Pages: 4, 5, 6, 8 |
| Results | Summary of main results | Report the mean values for the main categories of costs and outcomes of interest and summarise them in the most appropriate overall measure. | Pages: 3, 9, 10, 11 |
| Results | Effect of uncertainty | Describe how uncertainty about analytic judgments, inputs, or projections affect findings. Report the effect of choice of discount rate and time horizon, if applicable. | Pages: 4, 13, 14 |

| | | | |
|---|---|---|---|
| Results | Effect of engagement with patients and others affected by the study | Report on any difference patient/service recipient, general public, community, or stakeholder involvement made to the approach or findings of the study | N/A |
| Discussion | Study findings, limitations, generalisability, and current knowledge | Report key findings, limitations, ethical or equity considerations not captured, and how these could affect patients, policy, or practice. | Pages: 14,15 |
| Other relevant information | Source of funding | Describe how the study was funded and any role of the funder in the identification, design, conduct, and reporting of the analysis | Page 1 |
| Other relevant information | Conflicts of interest | Report authors conflicts of interest according to journal or International Committee of Medical Journal Editors requirements. | Page 1 |
